# Environmental surveillance for typhoidal *Salmonellas* in household and surface waters in Nepal identifies potential transmission pathways

**DOI:** 10.1101/2023.05.02.23289369

**Authors:** Christopher LeBoa, Sneha Shrestha, Jivan Shakya, Shiva Ram Naga, Sony Shrestha, Mudita Shakya, Alexander T. Yu, Rajeev Shrestha, Krista Vaidya, Nishan Katuwal, Kristen Aiemjoy, Isaac I. Bogoch, Christopher B. Uzzell, Denise O. Garrett, Stephen P. Luby, Jason R. Andrews, Dipesh Tamrakar

## Abstract

**Introduction:** *Salmonella* Typhi and *Salmonella* Paratyphi, fecal-oral transmitted bacterium have temporally and geographically heterogeneous pathways of transmission. Previous work in Kathmandu Nepal implicated stone waterspouts as a dominant transmission pathway after 77% of samples tested positive for *S*. Typhi and 70% for *S*. Paratyphi. Due to a falling water table, these spouts no longer provide drinking water, but typhoid fever persists, and the question of the disease’s dominant pathway of transmission remains unanswered.

**Methods:** We used environmental surveillance to detect *S*. Typhi and Paratyphi DNA from potential sources of transmission. We collected 1L drinking water samples from a population-based random sample of households between February and October 2019 Between November 2019 and July 2021, we collected monthly 50 mL river water samples from 19 sites along the rivers leading through the Kathmandu and Kavre Districts of Nepal. We processed drinking water samples using a single qpcr and processed river water samples using differential centrifugation and qPCR at time 0 and after 16 hours of culture enrichment. A 3-cycle threshold (Ct) decrease of *S*. Typhi or *S*. Paratyphi, pre- and post-enrichment, was used as evidence of growth. We also performed structured observations of human-environment interactions to understand pathways of potential exposure.

**Results:** Among 370 drinking water samples, *S*. Typhi was detected in 7 samples (1.8%) and *S*. Paratyphi A was detected in 4 (1.0%) samples. Among 381 river water samples, *S*. Typhi was detected in 171 (45%) and *S*. Paratyphi A was detected in 152 (42%) samples. Samples located upstream of the Kathmandu city center were positive for *S*. Typhi 12% of the time while samples from locations in and downstream the city had bacterial DNA detected 58% and 67% of the time respectively. Individuals were observed bathing in the rivers, washing clothes, and washing vegetables for sale in Kathmandu markets.

**Implications:** These results suggest that drinking water was not the dominant pathway of transmission of *S*. Typhi and *S*. Paratyphi A in the Kathmandu Valley in 2019. The high degree of river water contamination and its use for washing vegetables raises the possibility that river systems srepresent an important source of typhoid exposure in Kathmandu.

**Author Summary:** Understanding the dominant route of transmission of a pathogen is important for designing and implementing effective control strategies. Salmonella Typhi and Paratyphi which cause typhoid and paratyphoid fever infect an estimated 10 million people and kill more than 100,000 annually. In Kathmandu prior work suggested that stone spouts where people collected drinking water were contaminated and driving transmission of the diseases. However, many of these spouts no longer function, and people are still getting sick. We tested drinking water from households in this area as well as local river water and found that 13 drinking water samples were positive for S.Typhi and 15 were positive for S. Paratyphi and many river samples tested positive for these bacterium. River water samples were not often positive upstream of Kathmandu city center (12% positive for S.Typhi) but were often positive within the city center (58% positive for S.Typhi) and in rural areas up to 10 km downstream of the city (67% positive for S.Typhi). During sample collection, individuals were observed interacting with rivers by walking in them, washing clothes and washing vegetables for sale in markets. This study shows that drinking water may not be primary driver of typhoid transmission in the Kathmandu valley, but that sewage contaminated river water may be a foci of transmission into the wider population.

## Introduction

Typhoid fever, a fecal-oral transmitted febrile illness caused by *Salmonella enterica* serovars Typhi and Paratyphi A, B, and C, causes 10 to 20 million illnesses and over 100,00 deaths annually [1]. Improvements in the sanitation infrastructure in much of Europe and North America in the 20th century eliminated typhoid fever transmission in those areas[2]. However, typhoid fever remains highly endemic in many areas today, especially in Africa and South Asia [1]. Nepal, where this study takes place, has a high level of typhoid fever transmission with an estimated incidence rate of 330 cases per 100,000 person years and a seroincidence of 6.6 per 100 person years [1,3,4].

Currently inactivated whole cell (Ty21a) and oral vaccines (Vi) are used to prevent illness but these vaccines are only partially effective (∼55%) and are not given to children under two years of age, an especially vulnerable population [5,6].Antibiotics are used to treat infection but drug resistance, including extreme drug resistant strains, are increasing, rendering treatment more difficult [5,7].The World Health Organization (WHO) prequalified typhoid conjugate vaccine (TyVAC) generated much hope as a solution to ongoing transmission with phase three trials showing it to be 85% effective, but with few indirect effects of vaccination[8]. In a Bangladeshi TCV mass vaccination campaign, inoculated areas still had a high incidence of typhoid fever (96 cases per 100,000 person years) indicating that even effective vaccines alone cannot end transmission[8]. Ending transmission also must also address environmental and political issues.

Clarifying pathways of *S*. Typhi and *S*. Paratyphi transmission permits focused interventions to interrupt this transmission. Environmental surveillance has emerged as a pragmatic, low-cost, non-invasive approach to detecting different pathogens, including *S*. Typhi and *S*. Paratyphi, across different mediums to help identify modalities by which people are exposed[9–11]. Typhoid has been known to have different dominant mechanisms of transmission in different settings and at different time[12]. In some settings drinking water has been recognized as an important route of typhoid fever exposure, and a meta-analysis showed that people who drink water from unimproved sources have a greater risk of contracting typhoid fever[13].In Nepal, previous work implicated drinking water as a source of *Salmonella* exposure among the human population. They found evidence of frequent *S*. Typhi (77% sample positivity) and *S*. Paratyphi A (70%) DNA in water collected from public drinking water spouts, which led them to conclude “that the municipal water in Kathmandu is a predominant vehicle for the transmission of *S*. Typhi and *S*. Paratyphi A [in the Kathmandu Valley]”[9]. In the twelve years since their samples were collected, lowering aquifer levels and a large earthquake rendered many of the aquifers they tested inoperable yet typhoid cases in the Kathmandu Valley have remained high[3,14]. As such, other sources of transmission remain important to identify. A recent case-control study from Malawi found use of river water for cooking and cleaning to be the strongest individual risk factor for typhoid fever[15]. Additionally, researchers studying typhoid fever in Chile in the 1980’s had discovered typhoid fever causing bacteria in the river and irrigation water downstream of the capital, Santiago that was being used to irrigate crops[16]. This evidence was used to shape agricultural policy, banning the growth of salad greens downstream of Santiago and ultimately led to a decrease of typhoid fever incidence in the area[17]. The Kathmandu valley, a dense urban area with rivers running through it surrounded by agriculture, could potentially exhibit a similar cycle of typhoid fever transmission, but this exposure pathway has yet to be examined. This study seeks to understand the extent to which different water sources are contaminated with typhoid fever causing bacteria *S*.Typhi and *S*.Paratyphi A across the urban Kathmandu Valley and more rural Kavre district of Nepal. We test drinking water and surface water (river) samples from both Kathmandu and Kavre and record human river interactions to understand the potential role of other waterborne transmission routes of typhoid fever in this area.

## Methods

### Drinking water sampling

We leveraged a population-based household survey conducted between 2017 and 2019 in Kathmandu and Kavrepalanchok districts, which included urban, peri-urban and rural communities[18]. From that survey, we randomly sampled households for drinking water. At each household, team members collected a one-liter sample of the house’s drinking water in a sterile Whirlpak bag (Nasco product Number: **B01027)** and secured it with bag locking pipe closures (Nasco product number: **B01595**). Samples were either collected directly from the tap or poured into a Whirlpak bag. If the water container was too heavy to lift, the sample was spooned from the container into the Whirlpak bag, using the same cup or device that the household used to transfer the water. For households receiving water from a municipal water source, we collected the sample from the actively running household pipe inlet to the house. If water was stored before drinking, we collected the sample from its primary storage container (i.e. underground or above building water tanks). In cases where primary storage was inaccessible, we collected the sample from the family’s backup water reserve. For households using water from communal spigots, we collected water directly from the source spigot. If the communal spigot was not functioning, we collected a sample from a storage tank in the house, if the storage tank was inaccessible, sample was collected from water stored in a jug. For households purchasing water from a private vendor, in bottles, jars or tanks, we collected the sample directly from bottles or jars or primary storage jugs or tanks.

All samples were put on ice for transportation back to the laboratory. At each household questionnaires were collected on household drinking water sourcing, water purification processes using the Redcap Mobile application v5.19.15 (Vanderbilt University, Nashville,TN) on tablets.

### Limit of Detection and Spiking

We first characterized the sensitivity of the laboratory methods we utilized by performing a limit of detection assay on our proposed extraction and amplification processes. We used linearized standard plasmid containing *S*. Typhi STY0201 (132bp) and *S*. Paratyphi SSPA7038 (105bp) target gene for these experiments. We conducted a set of 11-10-fold serial dilutions starting at a concentration of 114 ng/μl to 114 *×* 10^-11 ng/μl for STY0201 and from 104 ng/μl to 104 *×* 10^-11 ng/μl for SSPA7038. Each assay was performed in 4 replicates. The mean Ct value and associated numbers of detected DNA copies were recorded for each dilution in excel.

We additionally conducted a series of spiking experiments to understand the limit of detection in settings that more simulated the samples that we collected of drinking and river water. We prepared a standard 0.5 McFarland standard inoculum (approx. 1.5×10^^8^CFU/ml) of Clinical *S*. Typhi strain in normal saline. We diluted the inoculum through a series of 8-10 fold dilutions with the most dilute solution having 1.5×10^0 CFU/ml *S*. Typhi. We then inoculated each dilution with 1.5×10^5 CFU/ml *S*. Typhi to 1.5×10^^0^ CFU/ml *S*. Typhi into separate 1L of commercially available mineral water bottles. These spiked water samples were mixed, then filtered through 0.45um Nalgene filter funnels. We performed DNA extraction on each spiked sample using Qiagen DNA PowerWater DNeasy Extraction Kit instructions and qPCR using the procedure available at (https://www.protocols.io/view/qpcr-protocol-cs9iwh4e).

### Drinking water assay

We stored water samples at 4°C and processed them within 24 hours of collection except on holidays. Processing involved filtering the sample through a 0.45µm cellulose nitrate filter in a 250ml Nalgene Analytical Test Filter Funnel using a vacuum pump (Shiva Industries, New Delhi, India). Using sterile forceps and scissors, we cut the filter membrane into 4 even pieces and inserted them into a 5ml Power Water DNA Bead Tube with the sample side facing inward. We performed DNA extraction using a Qiagen Power Water Kit (https://www.protocols.io/view/qiagen-dna-powerwater-dneasy-extraction-cs6ewhbe) After extraction, we performed real-time PCR (RT-PCR) to detect *S*. Typhi and *S*. Paratyphi DNA using previously published primers and probes from Nga et al. [19,20]. RT-PCR reactions were performed in 20 μl reaction volumes consisting of 10μl master mix (TaqMan Environmental Master Mix 2.0), 0.8 μl of the forward and reverse primers (10 μM concentration), 0.4 μl of the probe (10 μM concentration), 4 μL template DNA and 4μL DNase/ RNase-Free water, for 45 cycles. We ran all samples in duplicate, and included two nuclease free water PCR negative controls on each 96 well plate. We chose a cycle threshold (Ct) cutoff value of 39 for determining sample positivity, as this achieved 100% specificity with process controls.

### River water sampling

We collected monthly water samples between November 2019 and July 2021 from 19 environmental surveillance sites (ES sites) located at distances 1, 5 and 10 km upstream and downstream from the main river confluence point in the Kathmandu Valley and at three locations 5 km apart on the Punyamata river in the more rural Kavre District (Figure 3). We chose this sampling strategy because it allowed us to sample from a range of environmental settings, including locations upstream of the major city center where settlement was relatively sparse, concentrate the majority of sampling locations within the population dense areas, and sample at locations in agricultural areas downstream of the city. We collected each 50 mL river water sample directly into a Falcon tube, the exterior of which we then wiped down with a 0.5% bleach solution and 70% ethanol and placed into separate bags on ice for transportation to the laboratory. At the final sampling location of each day, research staff collected a field negative control by pouring sterile distilled water into a Falcon tube and placing it in the sample bag for transport back to the laboratory. At each site, team members collected information in the mobile application Redcap v5.19.15(Vanderbilt University, Nashville,TN) on abiotic factors that may affect bacterial input or survival in the water including observation of sewage pipes draining into the water, fecal matter, human / animal presence at the sampling pont, water pH, temperature, and oxygen reduction potential (Apera Instruments, Columbus, OH, AI311). Our field sampling protocol can be found online at (https://www.protocols.io/view/field-protocol-for-river-water-sampling-cs9hwh36)

### River water assay

Upon returning to the laboratory, we mixed samples by vortexing and pipetted each into three parts. We used a densitometer (DEN-1, Grant Instruments Ltd., England) on the first 5 mL of the sample to conduct a turbidity test. The second 200μl of sample was used to detect *Escherichia. coli* as a measure of fecal contamination. We created a 1:200 dilution of this sample aliquot in sterile PBS and then plated the solution onto 3M *E*.*coli/* Coliform Count Plates for overnight incubation at 37°C. Blue colonies with gas present the next day were recorded as *E. coli* coliforms.

We processed the remaining 45 mL of sample using differential centrifugation and enrichment PCR. We used different sample processing, extraction and assaying methods for the river water, compared with the drinking water samples, due to: 1) the different consistency, including a high level of particulate and sewage contamination in the former; and 2) the low PCR positivity in the drinking water survey. First, we centrifuged the sample at 2000 rpm for 1 minute to separate larger debris. We transferred the supernatant to a new sterile tube and then centrifuged it at 4000 rpm for an additional 25 minutes to obtain the bacterial pellet. We resuspended the pellet in 0.5ml of sterile distilled water and then pipetted it into 10ml of Selenite F enrichment media. We pipetted one mL of the Selenite F bacterial mixture out of the culture media before incubation, and performed DNA extraction using the Qiagen Blood and Tissue Kit manufacturer specifications. This sample was considered the time 0 hour extraction. The remaining 9.5mL of Selenite F broth was then incubated at 37°C for 16 hours. At the 16-hour time point, we performed DNA extraction on an additional 1mL of this liquid media following the same procedures, which we called the T-16 extraction. We stored DNA extracts at -20°C until further processing. We additionally incubated and processed an uninoculated Selenite F media control each day as a laboratory control.

We performed real-time PCR on the time 0h and time 16h extractions, in duplicate, using the same reaction volumes and conditions specifications described for the drinking water assay section (2.2), except the amplification for these samples was performed for 40 cycles instead of 45. Sample positivity was defined in two different ways. First, if either of the PCR duplicates from the time 16 h extraction had a Ct (cycle threshold) value of ≤ 35 the sample was considered positive. While other environmental surveillance studies have used Ct value cutoffs of up 38 or 39, we chose to use this more conservative measure to account for possible nonspecific binding of the primers and to ensure spurious results were not considered positive[21,22].We defined samples as potentially viable when the PCR results showed a minimum of 3 Ct downwards shift between extractions done at time 0 and time 16 and if the sample had a Ct value of < 35 at the endpoint (T16) extraction[21,22]. The river water laboratory processes can be found at dx.doi.org/10.17504/protocols.io.261ge3mxol47/v1)

### Statistical analyses

We assembled maps of drinking water locations and river sampling point contamination in R using a basemap created in Mapbox Studio[23,24].We obtained data on daily rainfall from weather station data obtained through the Nepal Department of Hydrology and Meteorology and plotted in R using *ggplot2 [25,26]*. Using a standardized protocol, hydrological catchments for each river sampling point were generated using the Arc Hydro extension in ArcMap 10.7 (ESRI, Redlands, California, USA)[27]. Catchments were then geospatially intersected with the WorldPop gridded population dataset for the reference year 2020 to estimate total upstream catchment population for each sampling location[28]. We modeled the proportion of samples as a function of distance from the river confluence point using generalized additive models with logistic link functions (R package “mcgv”)[29]. We conducted all statistical analyses using R (version 4.0.4). The code uses for cleaning the data and generating visualizations found in this text can be found at (https://github.com/chrisleboa/nepal_typhoid)

## Results

### Limit of detection results

In the limit of detection tests, the RT-PCR assay we used consistently detected samples down to the 10^−10^ dilution level, corresponding to a detection limit of 80 DNA copies per μl of sample. The average Ct value for *S*. Typhi DNA at this dilution was 35. At lower concentrations, the assay had lower sensitivity. For S. Paratyphi A, the minimum detectable dilution was also 10^−10^ which corresponds to 92 DNA copies per μl. The average Ct value for this dilution was 37. The minimum detectable dilution of spiked S. Typhi in 1 liter water samples was ∼1500 colony forming units per liter (3.17 log cfu/L) which resulted in an average Ct value of 36.

### Drinking water testing

Three hundred and seventy drinking water samples were collected between February and October, 2019. (146 in Kathmandu and 224 in Kavre) (Figure 1).

**Figure 1:**
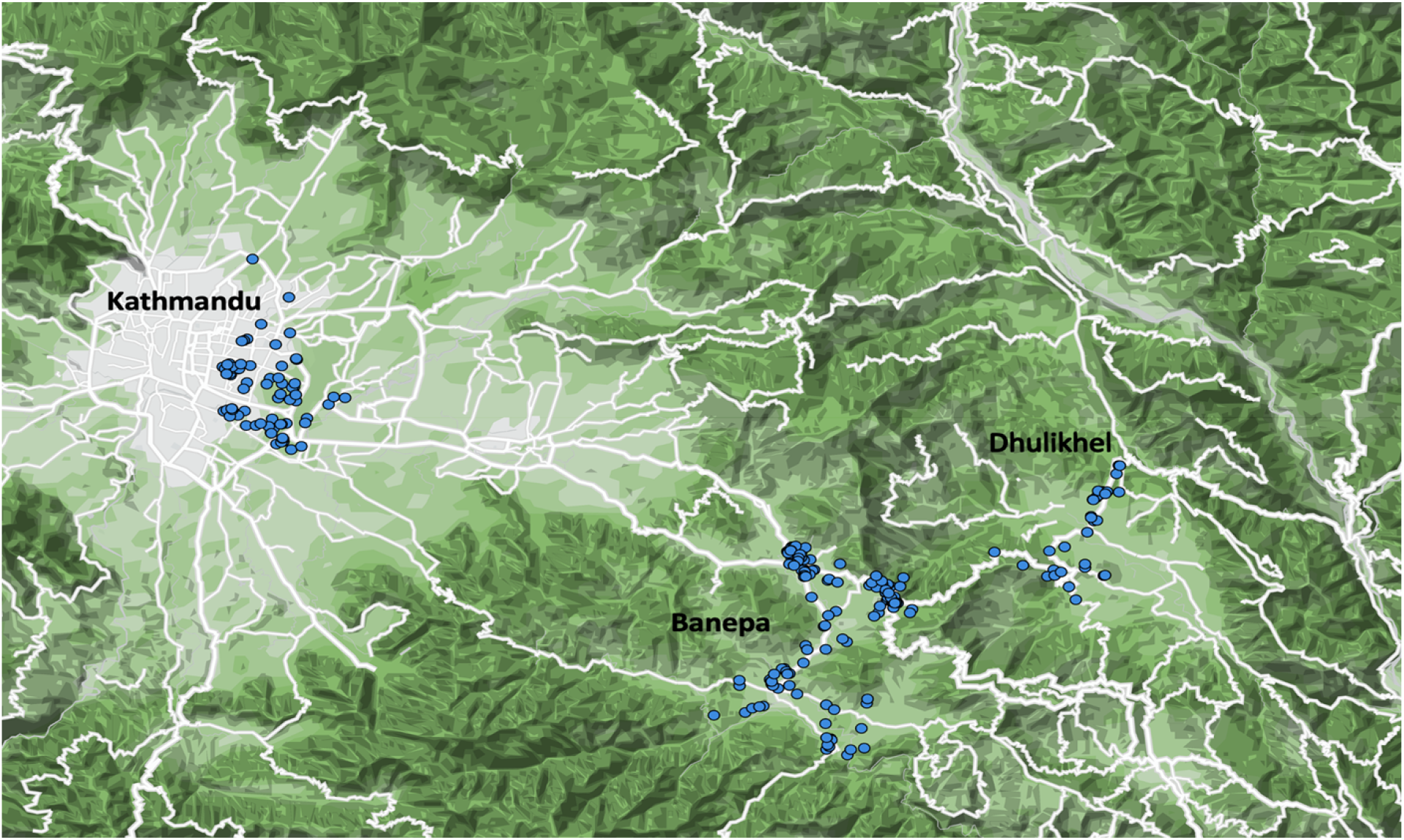
Drinking Water Sampling Locations. A random sample was drawn from a two-stage, cluster random sampling household survey of >25,000 households. The location of the 400 drinking water samples collected are shown as points. The aggregation of clusters of points in Kavrepalanchok reflects peri-urban areas where a higher number of households were sampled in the parent survey.

Households’ main source of drinking water varied: 154 samples (42%) came from the municipal water supply, 84 (23%) of the samples came from jugs purchased from private companies, 49 samples (13%) had an unknown origin, 41 samples (11%) came from surface water, 35 (9.5%) came from pipes of unknown origin, 5 samples (1.4%) were from a well at the house, and 2 samples (0.5%) came from water dropped off by a water tanker truck.

Households in the Kavre region were more likely to get water from the municipal water supply than those in Kathmandu (108/224 vs 46/146 p = 0.0014) and also were more likely to use surface water for drinking purposes (34/224 vs 7/146 p = 0.0019) while households in Kathmandu were more likely to purchase their drinking water from private companies, both jug and tanker samples (63/146 vs 20/224 p < 0.01). 77% of samples from pipes of unknown origin came from the more rural Kavre region. Of the 7 samples that tested positive for S. Typhi, one had come from Kathmandu while the other 6 originated from households in the Kavre region. Four of the positive samples came from municipal water (including the sample from Kathmandu, one came from a water sample from a private company, one came from surface water, and the last was undefined. Of the four samples that tested positive for S. Paratyphi, three came from Kathmandu and a single sample came from Kavre. Two of the samples’ origin was undefined, one came from a well and one came from the municipal supply.

### River water testing

We collected 381 water samples from the 19 sampling points, each site sampled 19 times between November 2019 and July 2021, on approximately a monthly basis, with brief interruptions due to COVID-19 associated lockdowns. Among these, 45% (n=171) tested positive for *S*. Typhi DNA and 40% (n = 152) of the samples tested positive for *S*. Paratyphi A at the time 16-hour extraction. Overall positivity was much lower before the enrichment step, with only 13 samples testing positive for *S*. Typhi (3.4%) and three samples from *S*. Paratyphi (0.7%). Of samples that tested positive at endline, 92% (158/171) of the *S*. Typhi detected samples showed a > 3 CT shift between baseline and endline extractions while 95% (145/152) of *S*. Paratyphi samples showed this shift, suggesting viability. Of all samples run, 42% (158/381) showed viability (a reduction of 3 Ct between time 0 and 16 extractions) for *S*.Typhi and 38% (145/381) showed potential viability for *S*. Paratyphi.

Sample positivity for typhoidal *Salmonellas* varied by month, with a lower percentage of locations testing positive during monsoon months (June - September) compared to the other parts of the year (S. Typhi 32% in summer, 52% non-summer p =0.012, S. Paratyphi 24% summer, 42% non-summer p = 0.025) (Figure 2). We observed a similar reduction in *E. coli* concentration during monsoon months.

**Figure 2.**
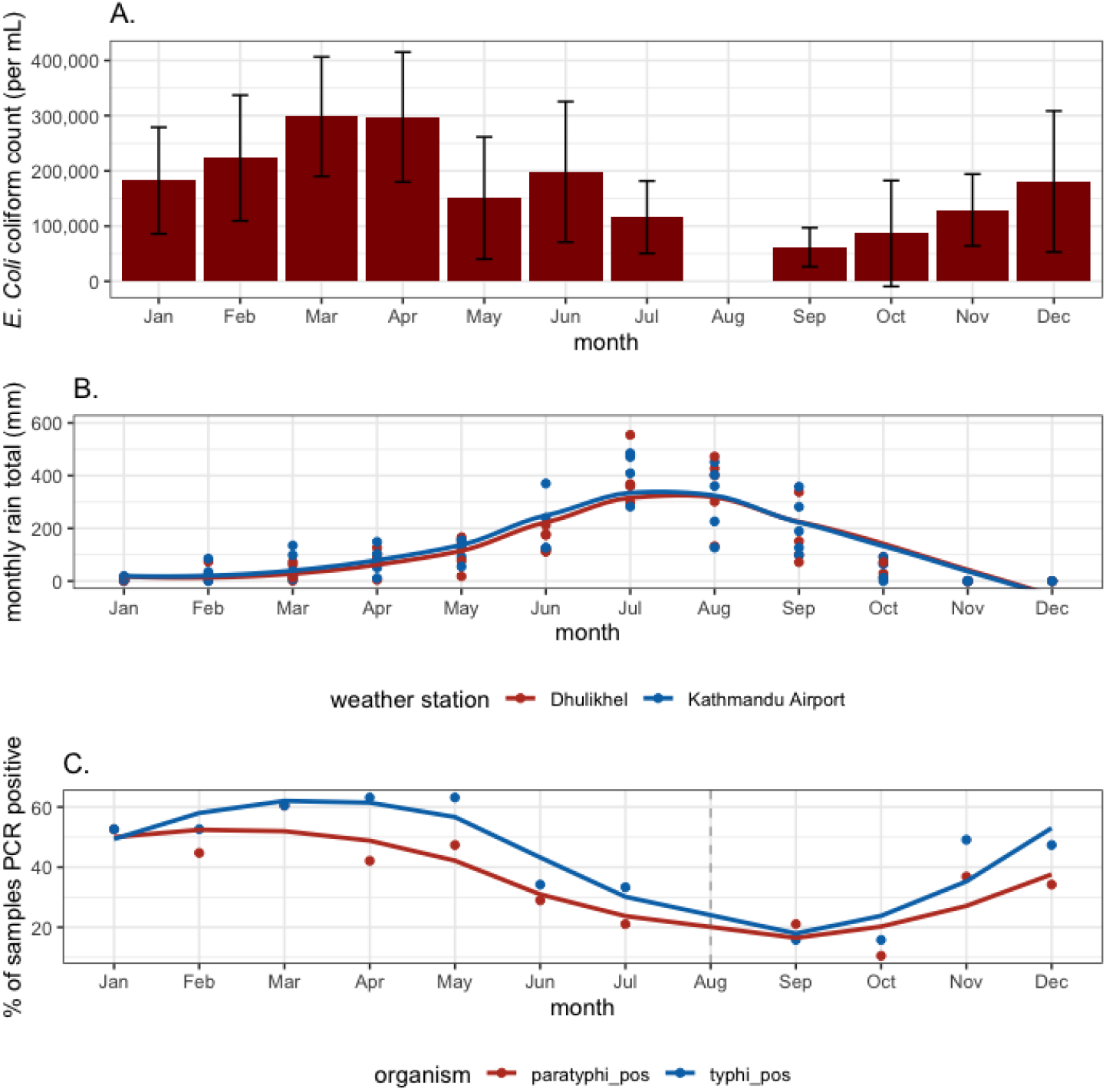
Presence of typhoidal *Salmonella* DNA in river water samples, *E. coli* concentration and rainfall by month. A. The median number of E. Coli coliforms present per mL of sample by month. Note August was missed in both years of sampling due to Coronavirus Lockdowns. B. Monthly rainfall in Kathmandu and Kavrepalanchok. C. The proportion of river water samples PCR positive for *Salmonella* Typhi or Paratyphi A by month.

River water contamination with typhoidal *Salmonellas* varied spatially. *S*. Typhi was detected at 10 sampling sites more than 10 times, representing over 50% of sampling trips while *S*. Paratyphi was detected at 9 sites on at least 10 occasions (Figure 3) Three sampling locations tested positive for *S*. Paratyphi once; two sites tested positive a single time for *S*. Typhi. Central Kathmandu (samples 1km and 5 km upstream of confluence) was highly contaminated with 61% (86/140) of samples testing positive for *S*. Typhi and 53% (74/140) positive for *S*. Paratyphi. Samples collected downstream of the city were positive 65% (39/60) of the time for *S*. Typhi and 60% (36/60) for *S*. Paratyphi A. Areas upstream from densely populated areas (samples 10 km upstream from the confluence) generally had lower positivity 13% (10/80) for S.Typhi; 6.3% (5/80) for *S*. Paratyphi. A chi square test with two degrees of freedom shows these groupings to be significantly different p <0.001 for both *S*. Typhi and *S*. Paratyphi. Similarly, samples collected from Kathmandu Valley were more often positive than those collected from the more rural Kavre region. For *S*. Typhi, 49% of Kathmandu Valley samples were positive (156/320) while 25% (15/60) of Kavre samples were positive. *S*. Paratyphi showed a similar pattern with 42% (135/320) and 28% (17/60) of samples testing positive in Kathmandu and Kavre respectively.

**Figure 3.**
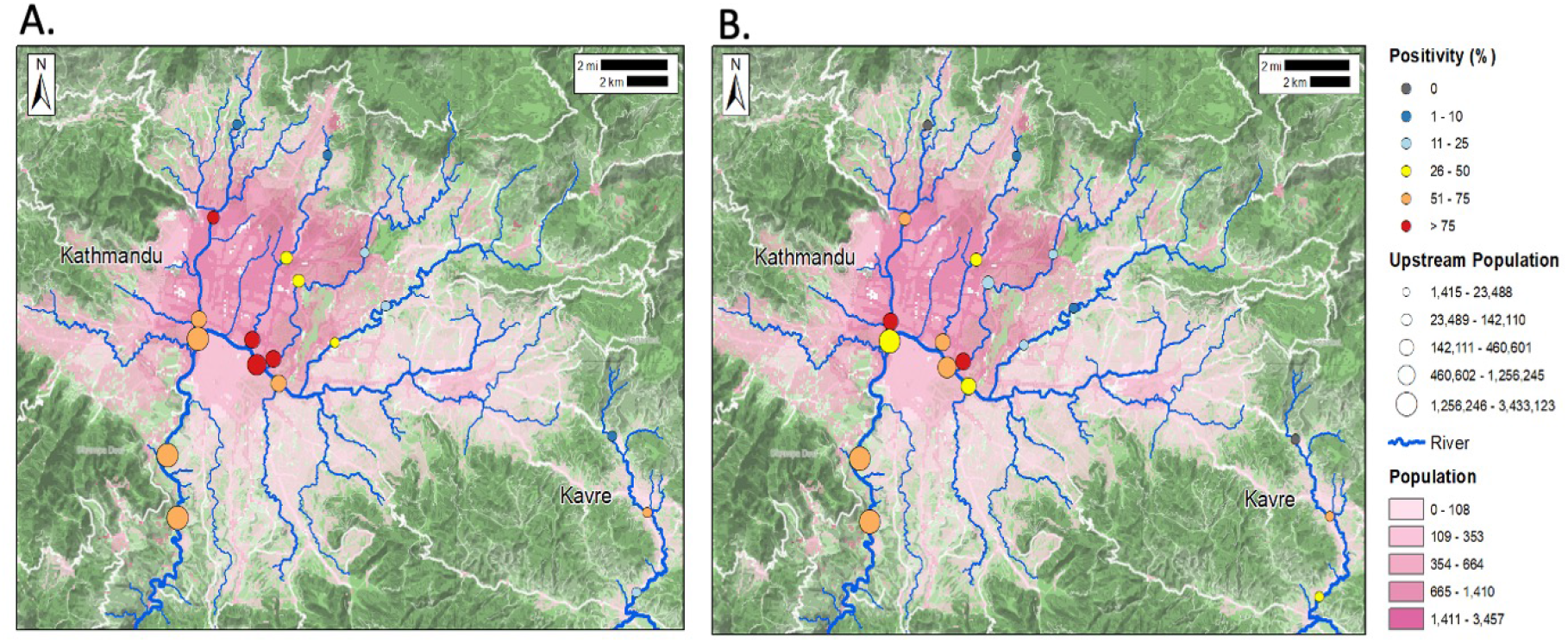
Frequency of detection of *Salmonella* Typhi and *Salmonella* Paratyphi A in river water by sampling location. Nineteen samples were collected from each of the 19 sampling points, and the proportion positive for *Salmonella* Typhi or *Salmonella* Paratyphi A are indicated by color of the sampling point. The size of the sampling point indicates the upstream population. Population density is indicated by shading.

### Observations of river usage

At each sampling location, we conducted observations of possible sources of fecal contamination of waterways and human interactions with the rivers to understand how these water sources may facilitate exposure to *S*. Typhi and *S*. Paratyphi. At fifteen of the nineteen sampling locations, there were sewage or rainwater runoff pipes entering the river system (13 from Kathmandu and 2 from Kavre). At 8 of 19 of these sites, we observed untreated sewage actively draining into the water. There were agricultural fields along the banks of the river at 7 of 19 of the sampling sites, including the two most downstream sampling locations from the Kathmandu valley. We observed in one location, an irrigation pipe leading from the river to a nearby field.

Our field team also observed people interacting with the rivers. People walked through the rivers at 7 of 19 sampling locations, washed clothes in the rivers at 8 of 19 sampling locations, bathed in the rivers at 3 of 19 sampling locations, and washed vegetables in the rivers two sampling points on twelve separate occasions (Figure 4). The corresponding water samples tested positive for S. Typhi on 9 occasions when people were observed walking, 5 occasions when people were washing clothes, four occasions when people were bathing and 4 occasions at which people were washing vegetables. Conversations with these individuals indicated that the vegetables were being washed before sale in markets in and around Kathmandu.

**Figure 4:**
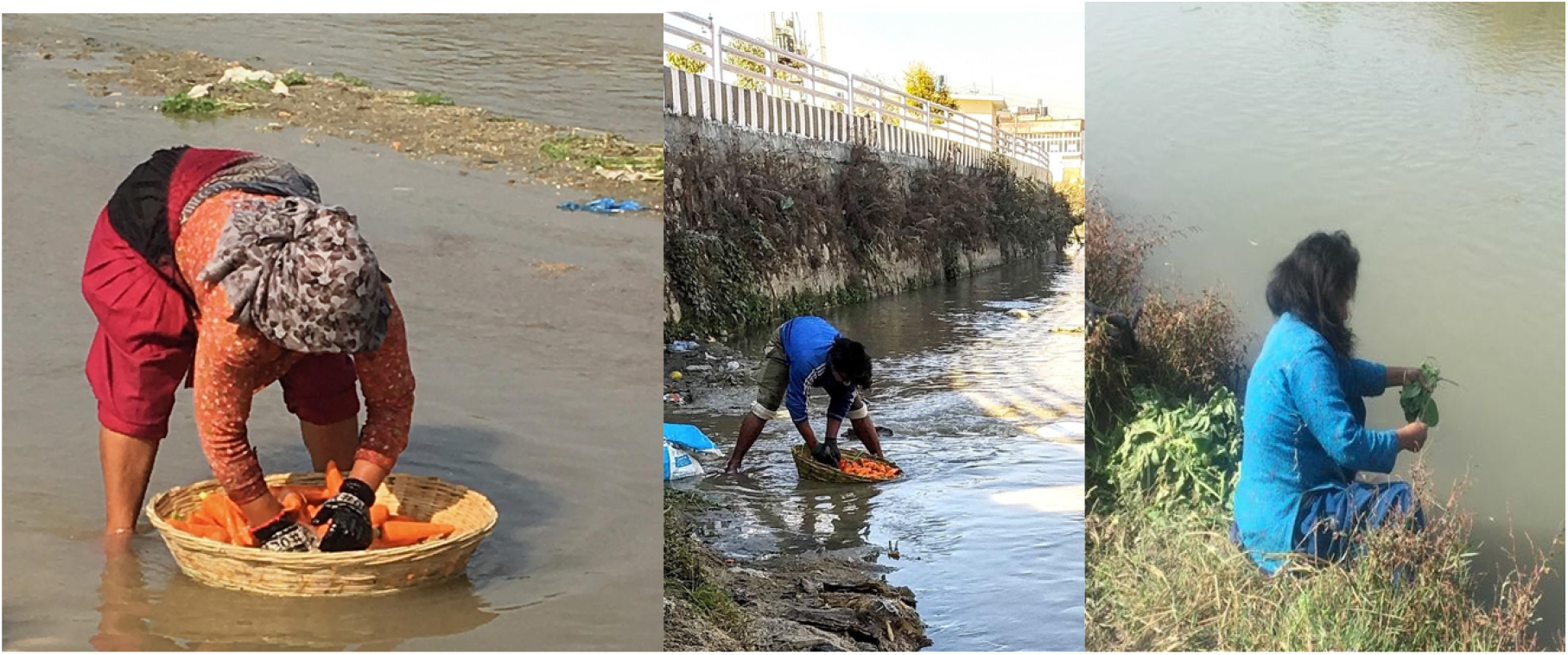
Observations of individuals washing vegetables: At two sampling points across twelve instances individuals were observed washing vegetables from the market. These images show that carrots and spinach were the most common vegetables being washed.

## Discussion

### Drinking water and fecal-oral transmission

In a population-based sample of household drinking water, we found low levels of contamination with typhoid fever causing *Salmonellas*. By contrast, sampling of river waters revealed the presence of *S*. Typhi and *S*. Paratyphi A in over 40% of river water samples.

Contamination with typhoidal *Salmonellas* occurs more frequently in river segments within and downstream of densely populated areas. Additionally, the use of sewage-containing river water for washing vegetables, and the potential uses of irrigation of nearby agricultural fields, suggests possible routes for direct exposure or the food-borne spread of typhoidal *Salmonellas*.

Our findings diverge from previous studies in South Asia, which have reported high rates of *S*. Typhi in drinking water and had suggested drinking water as the primary route for bacteria exposure [9,13]The Karkey et al study in Lalitpur, an area within the Kathmandu Valley, found 77% of the samples collected from 10 stone spouts contained *S*. Typhi DNA [9]. When we a representative sample of 370 hh from across the Kathmandu Valley and Kavre we found that no households reported using these spouts for drinking water procurement. Many of these spouts have become inoperable due to groundwater over extraction (the aquifer below Kathmandu is falling at a rate of 2 meters per year) and earthquake damage [14,30–32], Kathmandu suffers from increasing levels of municipal water scarcity, with demand (360,000 m^3^/day) exceeding supply (ranging between 90,000 and 150,000 m^3^/day) [30]. In our study, less than 50% of households sampled from Kathmandu or the Kavre region (154/370) used municipal water as their main drinking water source although slightly more households used municipal water from Kavre than did in Kathmandu[33]. In our study, private water companies (63/146) supplied much of Kathmandu’s water, collecting water from uncharacterized areas outside the valley and selling it by the truckload or in 20L bottles to city residents [33,34] The sources of private company water may be from less populated areas and have less sewage in the groundwater or surface water than water originating in the Kathmandu Valley. In Kavre, a significant portion of households used surface water or locally constructed pipe systems for their drinking water (54/224). As this region is not downstream from a large population center, these surface samples were less likely to be contaminated with sewage effluent before reaching these rural and semi-rural households. We found that 7 drinking water samples were positive for *Salmonella* Typhi and 4 of drinking water samples had detectable levels of *S*. Paratyphi A, five of the positive samples (45%) came from the municipal water supply which due to the sporadic allows for surrounding sewage to leech into the pipes[35,36]

### Surface water as a foci for transmission

River water yielded high rates of positivity (*S*. Typhi 45% and *S* Paratyphi 42%), particularly in segments located within and downstream from densely populated areas. These findings indicate fecal matter from population centers is entering river systems, and that the rivers are a potential hazard to human health even many kilometers past city boundaries. Our findings augment similar work describing high levels of fecal coliforms in the waters in Kathmandu’s river system[37]. In Kathmandu, municipal sewage systems are absent or incomplete[38]. Our findings indicate that the samples from downstream river locations, adjacent to agricultural fields, were positive for S.Typhi 65% of the time suggesting that nearby irrigation water may also be contaminated and used on crops. This potential pathway of transmission would mirror what was described in Chile in the 1980’s[16,17]. Additional analysis of irrigation water in areas downstream of Kathmandu could indicate whether irrigation of crops with S. Typhi contaminated water presents a potential foci of transmission. Our structured observations added evidence that rivers may be at locations at which people are exposed to typhoidal Salmonellas, as we observed agricultural workers washing carrots and spinach in the rivers before the market. Carrots especially are a potential source of contamination as they are commonly eaten raw in Nepal. Our work provides evidence that river water in Kathmandu is heavily contaminated with *S*. Typhi and Paratyphi, including several kilometers downstream of the population center and in some cases, people directly contact rivers or use river water for washing products. However, the scale to which this contact occurs is still not well characterized. The positivity of river water samples could simply be evidence of shedding from infected individuals and not a primary source of risk for disease exposure among the general population. Additional epidemiological investigations could be used to better understand the pathways of transmission in this context.

### Surface water seasonality

We found both *S*. Typhi and *S*. Paratyphi A contamination across the greatest number of sample sites during the dry season (October - May), we also found higher levels of *E. coli* contamination during these months. The higher levels of *S*. Typhi contamination during these months contrasts the seasonal pattern of typhoid fever incidence in the Kathmandu Valley, which have been reported to peak during the monsoon season [9,39]. We hypothesize the river may have more detectable levels of *S*.Typhi during dry seasons due to dilutional effects of monsoon rains increasing the overall water flow through the valley’s river systems while the levels of sewage generated remain constant.

### Environmental surveillance and case data

We found sample locations from the Kathmandu Valley more often contaminated with S. Typhi and Paratyphi (49% *S*. Typhi and 42% *S*. Paratyphi) compared to locations from the more rural Kavre District (25% *S*. Typhi and 28% *S*. Paratyphi). Sero-epidemiological data from these two regions similarly shows higher levels of *S*. Typhi exposure in Kathmandu (6.6 per 100 person years) compared to Kavre (5.8 per 100 person years)[4]. Environmental surveillance for pathogens can be used to not only show sources of transmission but as a sentinel surveillance tool to understand ongoing nearby disease transmission. Wastewater based epidemiology has been used in the COVID-19 pandemic to understand trends in otherwise unreported cases, but more work must be done to relate a positive environmental sample with associated disease incidence in nearby populations[11,40,41].

### Limitations and next steps

The results of this work must be interpreted in the context of numerous limitations. First, drinking water samples were analyzed without an enrichment step while river water samples were extracted both pre and post enrichment (the T0 and T16 samples). If an enrichment step was also added to drinking water samples, they may have more often shown detectable levels of *S*. Typhi and *S*. Paratyphi A. The earlier work by Karkey et. al also did not include an enrichment step before quantification through qPCR[9]. We attempted to culture and isolate *S*. Typhi from the water samples to directly demonstrate the viability of the organism in these samples, but were unsuccessful in doing so due to growth of other environmental bacterium, as was the experience of earlier work trying to culture S. Typhi from water[42]. To overcome this limitation, we used enrichment PCR in the river water samples, as previously described by the EPA for use on *Yersinia pestis* and other bioterrorism agents [21]. With this technique, a 2-cycle threshold shift represents a 4-fold increase in DNA detected, which is greater than would be expected by technical variability, providing evidence of bacterial growth but it is a proxy for the number of colony forming units present [34,43,44]. A positive river water sample test gives a binary, crude result indicating circulating disease nearby, but it is not possible to identify where the bacterium originated.

Additionally, the COVID-19 pandemic spurred lifestyle changes in the Kathmandu Valley, possibly shifting exposure pathways as well as overall typhoid fever incidence during this study period. Additional months of data collection could be used to see if the same trends identified in this publication are reproduced independent of potential influences of the pandemic.

## Conclusion

In population-based sampling of drinking water sources in the Kathmandu Valley, we found low abundance of typhoidal *Salmonella* DNA, in contrast to an earlier study. Drinking water samples that tested positive came from both the Kathmandu and the Kavre districts and came from municipal pipes as well as from a private jug sample, surface water sample and from a well. We found that rivers flowing through Kathmandu were highly contaminated with viable *S*. Typhi and *S*. Paratyphi A. These findings, together with observations of human use of river water for vegetable washing, suggest rivers as a potential transmission focus for typhoid fever in the Kathmandu Valley. Fecal contamination of river water should be addressed through improved sewage and sanitation infrastructure, and while such efforts are planned, they have yet to be implemented. A deeper understanding of the transmission pathways of typhoid fever in this region would frame effective measures for controlling its spread. These results show other sources beyond drinking water should be considered as possible pathways for transmission in this geographical context.

## Data Availability

The protocols used for this publication are available on protocols.io with links in the text. The code used for cleaning data, analysis and visualizations is availible on github and linked in the text and the data used for this publication has been uploaded to dryad https://doi.org/10.6078/D1ZD84.

## Abbreviations

DNA: Dideoxyribonucleic acid
qPCR: quantitative polymerase chain reaction

## Acknowledgements

A large team of individuals from multiple institutions helped make this project possible. Puspa Raj Bhatt, Bipin Thapa, Anil Khanal, Karuna Timilsina, Rojina Bhaila Shrestha, Natasha Shrestha, Shisir Ranjit, Lokmani Bhatt, Ashmita Karmacharya, Apekshya Chaulagain, Sudan Maharjan, Sabin Bikram Shahi, Sudichhya Tamrakar, Aastha Shrestha, Laxmi Chauguthi, Aarjya Tara Bajracharya, Suraj Jakibanzar, Melina Thapa and Neeru Suwal have helped with Sample/Data collection.

## Author Contributions

KA, DT, SL, CL, AY, SnehaS, and JA were responsible for project conceptualization. KA, KV, SnehaS, JA, CL, SN, and DT performed project coordination. JA, SL, KA and AY secured funding for this work. CL, JS, JA, KS, AY, SN, SonyS and SnehaS worked on methodology. JS, SnehaS, MS, SN, SnehaS, RS, CL, and many in the acknowledgements conducted the investigation. CL, KA, NK, JA, IB and KV procured study resources. CL, KA, AY, and CU worked on software and formal analysis. JA, DT, SL, NK, SN, RS and JS provided supervision. CL, JA and CU created visualizations. SonyS, SnehaS, JS and JA worked on validation. CL, SnehaS and JA wrote the original draft. All authors supported the reviewing and editing of this manuscript.

